# Leveraging Digitization, Archiving and Artificial Intelligence to Re-examine Predictors of Sustained Mental Health Care Engagement in Ugandan First-Episode Psychosis Patients: A Study Protocol

**DOI:** 10.64898/2026.06.02.26354672

**Authors:** Emmanuel Kiiza Mwesiga, Dickens Akena, Janet Nakigudde, Blessed Tabitha Aujo, Rossette Immy Ndigamanya, Angel Nanteza, Muyanga Mark, Andrea Kaddu Kaggwa, Sophia Balinga, Anne Nanyonga, Byamah Brian Mutamba, Rita Auma, Edith Kebirungi, Hafsa Lukwata, Phillip Pido Oyat, Wilber Ssembajjwe

**Affiliations:** Department of Psychiatry, College of Health Sciences, Makerere University, Kampala, Uganda; Psychosis Working Group, Department of Psychiatry, Makerere University, Kampala, Uganda; Mental Health Department, School of Medicine, Soroti University, Soroti, Uganda; Capacity Development Fellows of Digitizing Psychosis Study of the Mental Health Dataprize Africa; MRC/UVRI/LSHTM Uganda Research Unit, Kampala, Uganda; Butabika National Referral Mental Hospital, Kampala, Uganda; Heart to Heart Spaces, Uganda; YouBelong, Uganda; Mental Health Division, Ministry of Health, Uganda

**Keywords:** first-episode psychosis, mental health care engagement, digitization, archiving, artificial intelligence, natural language processing, Uganda, low- and middle-income countries

## Abstract

**Background:** We previously examined the burden and predictors of sustained mental health care engagement in Ugandan first episode psychosis patients by retrospective chart review methods. However, the extensive requirements of chart reviews meant that we could only extract data from a random 10% sample of 1,677 newly enrolled Ugandan first episode psychosis patients at Butabika National Referral Mental Hospital in 2018. The Hekima Platform has been designed to transform handwritten files into datasets for analysis.

**Objectives:** This study aims to: (1) utilize the Hekima Platform to transform paper-based clinical charts of all 1,677 Ugandan psychosis patients enrolled at Butabika Hospital for the first time in 2018 into a standardized, anonymized longitudinal database; and (2) re-examine predictors of sustained MHC engagement in this cohort.

**Methods:** We will digitize and archive all patient charts. We will then use the Hekima Platform to extract handwritten clinical data into machine-readable text using user-trained machine learning and deep learning models and natural language processing (NLP) techniques to generate a structured, anonymized database. A minimum 10% random sample of extracted data will be manually validated using Cohen’s kappa. For the analytical aim, descriptive statistics, bivariate analysis, and multivariable logistic regression will model predictors of sustained engagement, with exploratory machine learning approaches used as a complementary analytical strategy. Ethical approval has been obtained from the Uganda National Council for Science and Technology and Butabika Hospital’s Research Ethics Committee.

**Expected outcomes:** Patient clinical charts are a rich data source but there are extensive requirements to be able to use them for research. This study will generate the first AI-assisted, standardized longitudinal database from handwritten psychiatric records in Uganda, enabling well-powered analyses of predictors of MHC engagement. Findings will inform targeted interventions to improve retention in care and will offer a scalable model for mental health research in low- and middle-income countries.

## Introduction

Sustained engagement with mental health care after a first episode of psychosis is one of the strongest modifiable predictors of patient outcomes, including quality of life, disability, social functioning, and mortality (1, 2). Sustained engagement refers to not only coming to care but being involved in decision making. The determinants of engagement operate across multiple levels. They could be individual factors including illness symptoms, comorbidity, insight. They could be interpersonal especially through family involvement and stigma. Further, organizational factors like appointment systems, medication continuity, clinician– patient interaction must be considered. It is also important to consider community factors including distance and transport as well as policy factors like service organization and workforce distribution (3-5). In Uganda and similar settings, these factors are amplified by constrained service capacity and fragmented care pathways, including severe workforce shortages and limited system infrastructure for longitudinal follow-up (6, 7).

There is still limited data on the determinants of sustained mental health engagement in patients with a first episode of psychosis from low resource settings. At Butabika National Referral Mental Hospital (Butabika Hospital) in Uganda, we previously attempted to identify these factors. We hypothesized that thee determinants of are frequently documented, but not implicitly, within routine clinical notes (8). Among 1,667 patients presenting to the hospital with a first episode of psychosis (FEP) in 2018, a retrospective chart review was performed. However, the burden of the task made it unfeasible. Therefore, in a random sample of 167 files, we performed a retrospective analysis. We found that fewer than 20% engaged with the service more than twice in the subsequent year after initial admission (9). More concerning, 3.4% of patients who disengaged died within the following year, including deaths by suicide (9). Female sex and depressive symptoms were associated with sustained engagement, but the study was underpowered to assess a broader set of predictors and to examine multilevel causal pathways (4, 5).

Traditional research approaches have struggled to exploit the depth and breadth of information contained in patient clinical files at scale. Manual chart abstraction is slow, costly, and impractical for large cohorts, especially where records are predominantly handwritten and archived in analogue formats (10). As a result, the field has lacked sufficiently powered, context-rich datasets to quantify predictors of sustained engagement and to test causal hypotheses in African psychosis cohorts (3, 4). Digitization, archiving, and the adaptation of artificial intelligence (AI) methods for clinical chart data offer a feasible route to unlocking these underutilized records for research and service improvement. AI-enabled pipelines can extract variables from unstructured notes using natural language processing (NLP), harmonize inconsistent documentation into standardized fields, and scale the review of large volumes of files far beyond what is achievable by manual methods (11-15). Machine learning (ML) and deep learning (DL) approaches can iteratively improve extraction and harmonization as additional data are processed, supporting the creation of a living research database that remains relevant to clinical practice. Because implementation is as critical as technical performance, we will anchor development and rollout within the Consolidated Framework for Implementation Research (CFIR), explicitly attending to intervention characteristics, inner and outer setting constraints, user capabilities, and implementation processes to maximize feasibility, acceptability, and sustainability within Butabika Hospital and beyond.

For this study we aim to Leveraging Digitization, Archiving and Artificial Intelligence to Re-examine Predictors of Sustained Mental Health Care Engagement in Ugandan First-Episode Psychosis Patients. We hypothesize that AI tools can transform patient clinical files into analysable longitudinal datasets that enable a well-powered re-examination of predictors of sustained mental health care engagement among people with psychotic disorders. To operationalize this hypothesis, we will use the Hekima platform. Hekima is a purpose-built, low-resource–adapted digitization and data-integration pipeline designed to turn legacy handwritten clinical charts into a secure, de-identified, longitudinal research database. In practice, Hekima supports end-to-end chart capture (scanning and archival), standardized de-identification and record linkage, automated extraction of structured variables from free-text notes via OCR/handwritten text recognition and NLP, and iterative quality assurance through human-in-the-loop review and audit trails. It is designed to harmonize heterogeneous clinician documentation into analyzable fields, generate research-ready datasets at scale, and enable downstream statistical and machine-learning analyses while maintaining role-based access controls, governance, and ethical safeguards, with a pathway for integrating AI-extracted structured data into routine health information systems. The overall objective of this study is to leverage digitization, archiving, and AI applications in the Hekima Platform to (i) transform patient charts for 1,677 Ugandan psychosis patients enrolled for the first time at Butabika Hospital in 2018 into a standardized anonymized longitudinal database, and (ii) re-examine predictors of sustained mental healthcare engagement in this cohort, generating evidence to guide scalable interventions to reduce disengagement and prevent avoidable morbidity and mortality.

## Methods

### Study design

This is a retrospective cohort study involving secondary analysis of patient clinical charts. It comprises two sequential objectives: (i) a methodological and technological aim to construct a digitized, AI-processed database; and (ii) an analytical aim to re-examine predictors of sustained MHC engagement.

### Study setting

The study will be conducted at Butabika National Referral Mental Hospital, Uganda’s primary mental health referral center, a 600-bed hospital located 13 km east of Kampala (16). The hospital serves a broad national catchment area and has been the site of prior first-episode psychosis research. It houses the physical paper-based clinical charts that form the data source for this study.

### Study population

Patients in Uganda diagnosed with a psychotic disorder and receiving mental health care from Butabika hospital. We will include all patients who: (a) presented for the first time to Butabika Hospital in 2018 (17); and (b) received a diagnosis of a psychotic disorder from a qualified mental health worker. Patients with a comorbid substance use disorder were excluded.

### Power calculation

All 1,677 patient charts of individuals newly enrolled with a psychotic disorder at Butabika Hospital in 2018 will be analysed. With an expected sustained engagement rate of approximately 20%, this yields approximately 333 engaged and 1,334 disengaged patients. Power calculations confirm that at α = 0.05, the study has over 90% power to detect odds ratios as small as 1.3 for common predictors of engagement, and maintains ≥80% power for less frequent predictors.

### Objective 1: Transforming patient charts into a standardized, anonymized longitudinal database

#### Digitization and archiving

All 1,677 paper-based patient charts will be scanned to create a digital archive housed on a secure, on-premise server. A document management system will be used to organize and catalogue the digitized files. Given the sensitive nature of psychiatric records, a bioethicist and data privacy experts will oversee all digitization procedures to ensure compliance with local and international data protection regulations. Service users will participate as data safety monitors throughout this process.

#### OCR-based text extraction and model training

Scanners incorporate OCR software capable of converting scanned images, including handwritten text into machine-readable text. To improve recognition accuracy for Ugandan clinical handwriting, we will collect standardized handwriting samples from current clinical staff at Butabika Hospital. Clinicians will write the sentence “THE QUICK BROWN FOX JUMPS OVER THE LAZY DOG” (covering all letters of the English alphabet), the digits 0–9, and common medical abbreviations (e.g., ‘od’, ‘bd’, ‘IM’). These samples will be used to train user-specific handwriting recognition models within the Transkribus platform.

Pre-processing steps will include image quality enhancement, annotation of a labelled training dataset, and iterative development and cross-validation of machine learning and deep learning models. For records of former staff whose handwriting samples are unavailable, manual expert verification will be performed.

NLP-based data structuring: Extracted machine-readable text will be processed using NLP techniques including tokenization (splitting text into individual words or tokens), stop word removal (removing non-informative words, including patient identifiers), and lemmatization (reducing words to root forms for consistency). This will generate a structured, standardized, anonymized longitudinal database suitable for statistical analysis (18, 19).

#### Data validation

Sensitivity analyses will compare machine-extracted data against variables manually collected in the previous study to assess consistency and determine the accuracy of the AI pipeline.

### Objective 2: Re-examining predictors of sustained MHC engagement

Using the anonymized database, lived experts for psychotic disorders and multidisciplinary clinicians will collaboratively define the keywords and phrases to be used in the machine learning model to capture individual, interpersonal, organizational, community, and policy-level predictors of sustained MHC engagement. The model will extract and standardize these terms across all participant records.

Data analysis: Descriptive statistics and bivariate analyses will characterize the study population and examine unadjusted associations between candidate predictors and sustained MHC engagement (defined as ≥2 clinical contacts in the two years following enrolment). Multivariable logistic regression will identify independent predictors of sustained engagement, adjusting for confounders. Where appropriate, complementary machine learning approaches (e.g., random forest classifiers) will be used for exploratory prediction modelling. Model performance will be evaluated using cross-validation and reported with metrics including precision, recall, and area under the receiver-operating characteristic curve (AUC). A pilot phase using approximately 100 charts will precede full implementation to assess AI performance, user acceptability, and workflow feasibility.

#### Data quality assurance

A multi-step quality assurance process will minimize risks associated with OCR errors and illegible handwriting. A minimum 10% random sample of OCR-extracted data will be manually reviewed against original charts, with concordance assessed using Cohen’s kappa(20). Previously validated datasets from the prior study will serve as benchmarks for extraction accuracy. Discrepancies will inform model retraining cycles. Quality assurance logs will document model accuracy, error rates, and correction steps throughout. All data will be stored on secure servers with access controls, encryption, audit trails, and version control. A data integrity review team and a designated data protection expert will provide oversight. De-identification protocols will be strictly enforced prior to any analytical use of the data.

#### Ethical considerations

The study has received ethical approval from the Butabika National Referral Hospital Research and Ethics Committee (BNRMH-REC-2025-5), the Uganda National Council of Science and Technology (HS6912ES) as well as institutional approval from the Butabika Hospital Administration. The proposal has also been presented to the Technical Working Group on Non-Communicable Diseases at the Ministry of Health. Data extraction from physical records will be conducted exclusively by Butabika Hospital records staff, with all outputs anonymized prior to transfer for analysis. Advanced encryption and secure storage will be employed throughout.

#### Lived experience involvement

We are working with three key groups. First are patient representatives from Butabika Hospitals peer support workers. Peer support workers are a key component of Butabika hospital. They are stable patients who work side by side with the clinical staff to disseminate information, make patient grievances known and support recovery of other patients. Second, Heart to Heart Spaces is a community-based mental health organization in Kampala, Uganda dedicated to creating safe, supportive and inclusive environments where individuals can openly navigate their mental health journeys. The organization works to promote mental wellness through advocacy, education and community initiatives that reduce stigma and encourage healing. Through safe spaces, mental health awareness programs, and community outreach, Heart to Heart Spaces supports individuals and families affected by mental health challenges. It also connects people to professional care by providing mental health education, referrals to therapists and psychiatrists and resources that make support more accessible and affordable. Heart to Heart Spaces and peer support workers will serve in a data safety monitoring role, overseeing procedures to safeguard patient rights and ensure cultural and ethical sensitivity in the handling of sensitive mental health records. They will also support analysis of the data in by identifying drivers for engagement to be examined by AI models.

YouBelong Uganda (YBU) is a Ugandan NGO that has been working since 2016 to shift mental health care away from hospitals and institutions toward family and community-based settings, with a focus on people living with Mental, Neurological, and Substance use (MNS) disorders (21). Their work spans mental health promotion, child protection, social inclusion, gender equality, and research, all aimed at building interconnected support systems suited to low-resource environments. A core part of their work involves helping patients discharge from Butabika Hospital and reintegrate into their families through psychoeducation an approach that has reached over 800 families while their more recent Wellcome UK-funded SCAPE-U study collaborates with people who have lived experience of psychosis to strengthen care across primary, community, and family levels. YouBelong Uganda, will conduct an independent assessment of our protocol to review our methods and see how well we met our proposed study.

#### Patient and public involvement

Butabika Hospital’s established Community Advisory Board (CAB), comprising health workers, caregivers, academics, and patients with lived experience, will participate as data safety monitors. Stakeholders including policymakers and clinicians will be invited to contribute to feedback sessions, ensuring that the digitization pipeline and AI tools developed are contextually appropriate, acceptable to users, and scalable beyond the study setting.

#### Dissemination

Study findings will be disseminated through peer-reviewed publications and presentations at relevant national and international conferences. Results will be shared with Butabika Hospital, the Ministry of Health Uganda, Makerere University, and community-based mental health advocacy groups. Dissemination will be designed to inform national strategies for improving mental health service delivery and to support policy dialogue on the ethical use of digitized health data in Uganda and similar settings.

### Study flow diagram

Figure 2 summarizes the overall study flow and shows how the two objectives are linked. The study begins with all eligible patients with first-episode psychosis newly enrolled at Butabika National Referral Mental Hospital in 2018. Objective 1 focuses on transforming the original paper-based clinical charts into a secure, standardized, anonymized longitudinal database through sequential digitization, archiving, OCR and handwriting recognition, NLP-based data structuring, de-identification, and manual validation against original records. This database then becomes the foundation for Objective 2, in which clinicians and lived-experience experts help define candidate predictors of sustained mental health care engagement before descriptive, bivariate, multivariable regression, and exploratory machine-learning analyses are conducted. Across all stages, the workflow is supported by ethics approval, data protection procedures, quality assurance, audit trails, and lived-experience oversight.

**Figure 1:**
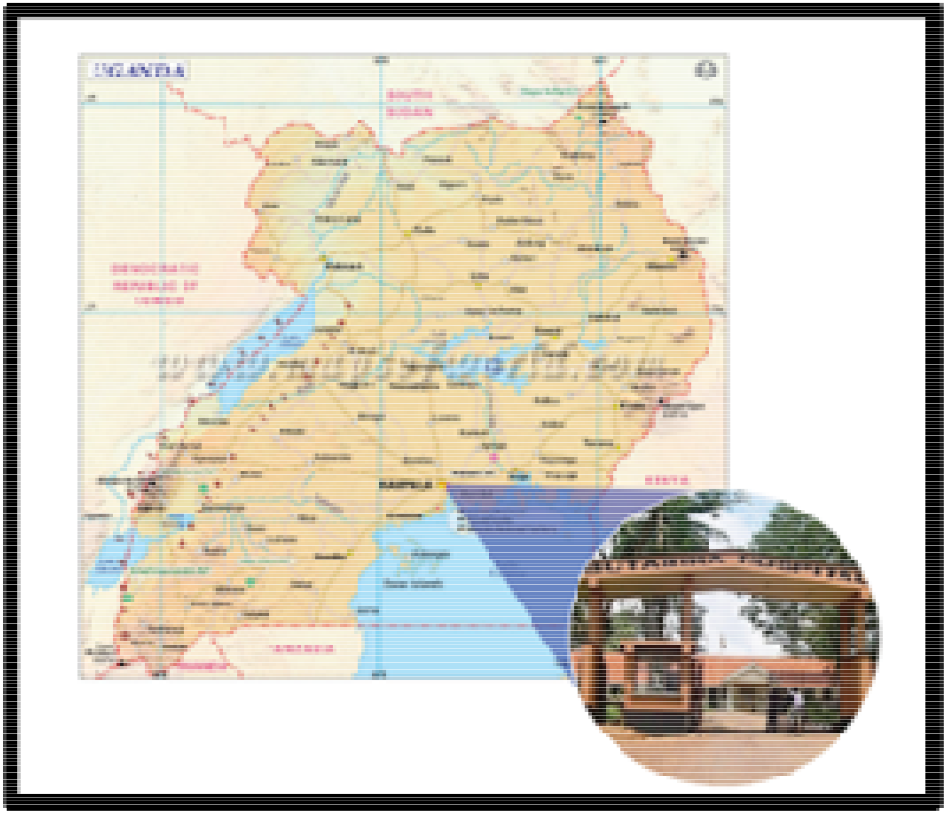
Map of Uganda with insert of Butabika Hospital

**Figure 2:**
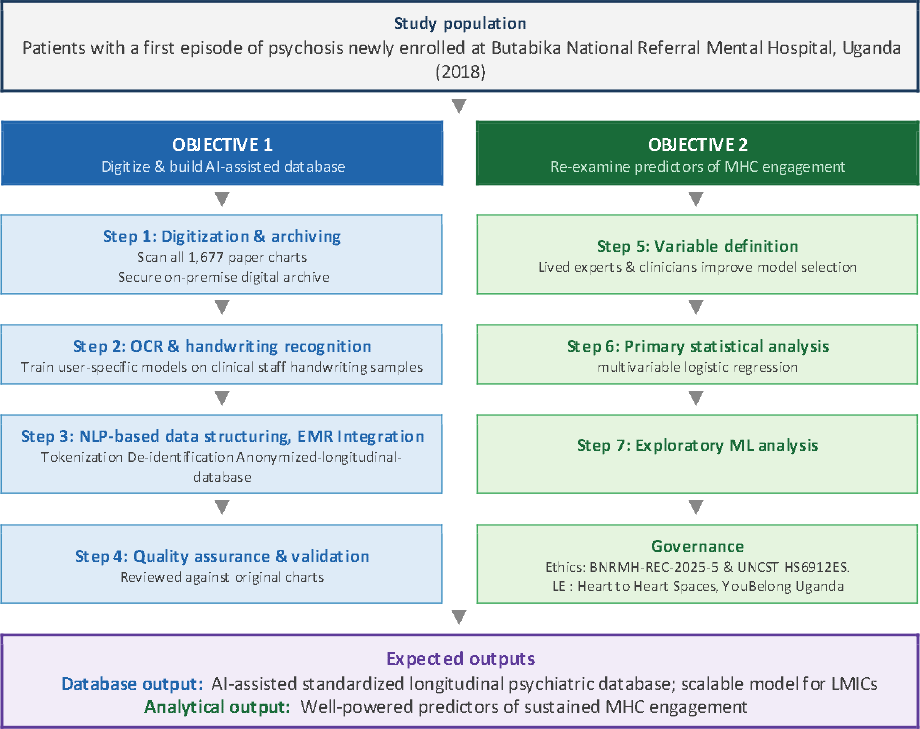
Central logic of the study objectives

#### Expected Outcomes and Significance

This study will generate the first AI-assisted, standardized longitudinal database derived from handwritten psychiatric clinical charts in Uganda. This database will enable well-powered analyses of the individual, interpersonal, organizational, community, and policy-level predictors of sustained MHC engagement among patients with first-episode psychosis in a low-resource setting. Identification of modifiable predictors of engagement will inform the development of targeted interventions to improve retention in mental health care, with direct implications for patient outcomes in Uganda and comparable settings across sub-Saharan Africa. The digitization and AI pipeline developed through this work will offer a scalable, ethical, and technically validated model for transforming handwritten records in resource-constrained health systems, with potential application beyond psychiatry. The study will also build institutional capacity in digital health and data science at Butabika Hospital and Makerere University, and will contribute to national dialogue on the ethical and policy dimensions of digitizing sensitive health records in Uganda.

## Data Availability

No datasets are associated with this protocol article. Data generated during the study will be made available in an approved repository in accordance with Wellcome Open Research data policy upon completion of the study subject to ethical constraints around patient confidentiality.

## Data Availability

No datasets are associated with this protocol article. Data generated during the study will be made available in an approved repository in accordance with Wellcome Open Research’s data policy upon completion of the study, subject to ethical constraints around patient confidentiality.

## Acknowledgements

The authors thank Butabika National Referral Mental Hospital, its clinical and records staff, and the Community Advisory Board for their support of this work. We also thank the lived experience experts and community stakeholders who have contributed to shaping the study design and oversight framework.

